# Octreotide may improve pharyngocutaneous fistula healing through downregulation of cystatins: a pilot study

**DOI:** 10.1101/2022.04.24.22273652

**Authors:** Jonathan Cohen, William Reed, Matthew W. Foster, Russel Kahmke, Daniel Rocke, Tammara Watts, Liana Puscas, Trinitia Cannon, Walter T. Lee

## Abstract

**Objective:** Pharyngocutaneous fistula (PCF) and salivary leaks are well known complications of head and neck surgery. The medical management of PCF has included the use of octreotide without a well-defined understanding of its therapeutic mechanism. We hypothesized that octreotide induces alterations in the saliva proteome and that these alterations may provide insight into the mechanism of action underlying improved PCF healing. We undertook an exploratory pilot study in healthy controls that involved collecting saliva before and after a subcutaneous injection of octreotide and performing proteomic analysis to determine the effects of octreotide.

**Materials and Methods:** Four healthy adult participants provided saliva samples before and after subcutaneous injection of octreotide. A mass-spectrometry based workflow optimized for the quantitative proteomic analysis of biofluids was then employed to analyze changes in salivary protein abundance after octreotide administration.

**Results:** There were 3,076 human, 332 S. mitis, 102 G. haemolyans and 42 G. adiacens protein groups quantified in saliva samples. A paired statistical analysis was performed using the generalized linear model (glm) function in edgeR. There were and ∼300 proteins that had a p<0.05 between the pre-and post-octreotide groups ∼50 proteins with an FDR-corrected p<0.05 between pre-and post-groups. These results were visualized using a volcano plot after filtering on proteins quantified by 2 more or unique precursors. Both human and bacterial proteins were among the proteins altered by octreotide treatment. Notably, four isoforms of the human cystatins, belonging to a family of cysteine proteases, that had significantly lower abundance after treatment.

**Conclusion:** This pilot study demonstrated octreotide-induced downregulation of cystatins. By downregulation of cystatins in the saliva, there is decreased inhibition of cysteine proteases such as Cathepsin S. This results in increased cysteine protease activity that has been linked to enhanced angiogenic response, cell proliferation and migration that have resulted in improved wound healing. These insights provide first steps at furthering our understanding of octreotide’s effects on saliva and reports of improved PCF healing.

## Introduction

Pharyngocutaneous fistulas (PCF) and salivary leaks are well known complications of head and neck surgery, occurring in up to 25% cases involving pharyngeal and laryngeal defects^1-3^ with the highest rates in those patients with prior radiation, chemotherapy, malnutrition, infection, and hypothyroidism.^4^ Management of these complications can be quite challenging for both patient and surgeon. Conservative management and observation success rates have been shown to be as high as 65%, decreasing to 30-40% in previously irradiated patients.^5^ Approximately 40% of those failing conservative therapies ultimately require surgical management, with even higher rates of surgical intervention in those with prior radiation.^6^ Beyond the medical and surgical implications, these complications often necessitate hospital re-admissions, prolong length of stay following surgery, and increase healthcare costs.

Saliva’s effects on wound healing have been a subject of study for many years, particularly based on the rapid healing of oral mucosa in comparison to skin wounds, much of which has been attributed to the presence of saliva and its proteins and peptides.^7-9^ In contrast, the role of saliva in wound healing in Head and Neck Cancer is less understood—as those afflicted with PCF experience delayed healing.^6,10^

The management of PCF has included the use of the octreotide, the first FDA approved synthetic analog of somatostatin. Somatostatin analogs have a number of physiologic effects, inhibiting both endocrine and exocrine secretions including growth hormone, prolactin, thyrotropin and amylase, as well as the reduced absorption of glucose, fat, and amino acids.^11^ However, its use in the setting of salivary fistulas has not been well studied. There have been anecdotal and case reports supporting the use of octreotide in the setting of PCF^12,13^ that have justified a placebo controlled clinical investigation of octreotide in PCF closure in Israel^14^.

We hypothesized that octreotide induces alterations in the saliva proteome, and these alterations may provide insight into the mechanism of action underlying improved PCF healing. We undertook an exploratory pilot study in healthy controls that involved collecting saliva before and after a subcutaneous injection of octreotide. Using a mass spectrometry-based workflow optimized for proteomic analysis of biofluids,^15,16^ we report these novel observations in salivary proteome alterations after octreotide administration.

## MATERIALS AND METHODS

This pilot study was approved by the Duke University Institutional Review Board. Healthy, adult participants provided a sample of saliva five minutes after rinsing their mouth with tap water and a two hour fast. The subjects were then given 100 µg octreotide in 1cc saline (Novartis) via subcutaneous injection. A second sample of saliva was then collected after 45-55 minutes. This interval was selected because peak plasma concentrations have been found to occur 0.4 hours after dosing.^17^ A total of four subjects were recruited, each providing a set of “pre” and “post” samples.

### Sample preparation

Samples were frozen at -80 °C and processed in a single batch. Samples were thawed and sonicated, and 100 μL of each sample was diluted 1:1 with 5% sodium dodecyl sulfate (SDS) in 50 mM triethylammonium bicarbonate, pH 8.5 (TEAB) followed by additional probe sonication and brief heating at 80 °C for 5 min. 100 μgs of each sample was adjusted to 75 μL with 5% SDS/TEAB. Samples were reduced by addition of 7.5 μL of 110 mM dithiothreitol and heated at 80 °C for 15 min. After cooling, samples were alkylated by addition of 7.5 μL of 250 mM iodoacetamide and incubation in the dark for 30 min. Finally, 9 μL of 12% phosphoric acid was added followed by 600 μL of 90% (v/v) methanol/100 mM TEAB, and the samples were processed using S-traps micro devices (Protifi). Samples were digested with 7 μg of Sequencing-Grade Modified Trypsin (Promega) at 47 °C for 2 h. Eluted peptides were lyophilized and reconstituted in 33 μL of 1/2/97 (v/v/v) trifluoroacetic acid/acetonitrile/water with incubation in a bath sonicator, followed by centrifugation. A study pool quality control (SPQC) sample was prepared by mixing equal quantities of all samples.

### Quantitative LC-MS/MS using data-independent acquisition

Thirteen microliters of the SPQC sample and 15 μL of individual samples were analyzed by microflow-liquid chromatography hyphenated with tandem mass spectrometry (LC-MS/MS) using an ACQUITY UPLC (Waters) interfaced to an Exploris 480 high resolution tandem mass spectrometer (Thermo). After direct injection, peptides were separated on a 1 mm x 10 mm 1.7 μm CSH C18 column (Waters) using a flow rate of 100 μL/min, a column temperature of 55 °C and a gradient using 0.1% (v/v) formic acid (FA) in H_2_O (mobile phase A) and 0.1% (v/v) FA in acetonitrile (mobile phase B) @ 100 μL/min as follows: 0-60 min, 3-28% B; 60-60.5 min, 28-90% B; 60.5-62.5 min, 90% B; 62.5-63 min, 90-3% B; 63-67 min, 3% B. A zero dead volume Peek tee (Thermo) was used post-column to introduce a solution of 50% (v/v) dimethyl sulfoxide/acetonitrile at 6 μL per min. The LC was interfaced to the MS with an Optamax NG ion source under heated electrospray ionization conditions with following tune parameters: sheath gas, 32; aux gas, 5; spray voltage, 3.5 kV; capillary temperature, 275 °C; aux gas heater temp, 125 °C.

The data-independent acquisition (DIA) analysis used a staggered, overlapping window method^18^ with a 60,000 resolution precursor ion (MS1) scan from 390-1020 m/z, AGC target of 1000% and maximum injection time (IT) of 60 ms and RF lens of 40%; data was collected in centroid mode. MS/MS was performed using targeted (tMS2) method with default charge state = 3, 15,000 resolution, automatic gain control (AGC) target of 1000%, maximum IT of 22 ms, and a normalized collision energe (NCE) of 30; data was collected in centroid mode. The DIA windows were generated using EncyclopeDIA (https://bitbucket.org/searleb/encyclopedia/)^19^ with a mass range of 400-1000 and 77 × 16 m/z windows, with a 36 window cycle. The MS cycle time was 1.6 s, and the total injection-to-injection time was 67 min.

### Quantitative analysis of DIA data

Raw MS data was demultiplexed and converted to *.htrms format using HTRMS converter (Biognosys) and processed in Spectronaut 15.5 (Biognosys). A spectral library was built using Direct-DIA searches of all individual files. Searches used a Swissprot database with *homo sapiens* taxonomy (downloaded on 10/16/20) and appended with a concatemer containing single amino acid variant peptides, as well as porcine trypsin. Protein sequences from *Streptococcus mitis, Gamella haemolysans* and *Granulicatella adiacens* were also downloaded from Uniprot and appended to generate a combined database with 25,944 entries. Search settings included N-terminal semi-tryptic specificity, up to 2 missed cleavages, fixed carbamidomethyl(Cys) and variable acetyl(protein-N-terminus) and oxidation(Met) modifications.

For DIA analysis, default extraction, calibration, identification and protein inference settings were used. iRT profiling was utilized to quantify precursors that did meet a q-value cut-off of <0.01 in a particular run. Protein quantification was performed at MS2 level using the MaxLFQ algorithm^18,20^, q-value percentile 0.2 settings (all precursors that passed a q-value in at least 20% of runs were included) with run-wise imputing, and local normalization^21^ using precursors that met a q-value in all runs (q-value complete).

### Statistical analysis

Statistical analysis used the generalized linear model (glm) function in edgeR,^22,23^ and data was visualized using ggVolcanoR (https://ggvolcanor.erc.monash.edu/).^24^

### Data availability

The raw mass spectrometry data, and data supplement, have been uploaded to the MassIVE repository (massive.ucsd.edu) with dataset identifier MSV000089053 and can be downloaded at ftp://MSV000089053@massive.ucsd.edu with password octreotide.

## RESULTS

### Quantitative proteomics of saliva from healthy controls treated with octreotide

We enrolled four healthy control subjects in a pilot trial to assess the proteome changes in response to administration of a single subcutaneous injection of octreotide. Saliva samples were obtained just prior to, and 45-55 min after injection and stored at -80 °C. Upon thawing, samples were probe sonicated and diluted 1:1 with SDS buffer followed by an additional round of sonication and a protein assay. The concentration and volume of samples from subject 1 were insufficient based on our target of >100 µL saliva and >100 µg of protein per sample (**Table S1**), so we proceeded with trypsin digestion from the three remaining subject samples using a commercial suspension-trapping (S-trap) devicei. The remaining samples were adequate.

Saliva proteomes were analyzed using microflow liquid chromatography (1 mm internal diameter x 100 mm length; 100 µL/min flow rate and 60 min gradient), post-column infusion of DMSO,^25^ and data-independent acquisition (DIA) using a staggered, overlapping window configuration.^18,19^ To compensate for the narrow peak widths at these higher flow rates, the DIA method used 16 m/z windows (8 m/z effective after deconvolution). Based on total ion current (**Figure 1**), slightly less than half of recovered peptides (15 µL out of 33 µL) were analyzed in singlicate in a block-randomized run order (**Table S1**). A study pool quality control (SPQC) sample, containing an equal mixture of each sample, were analyzed in triplicate at the beginning, middle and end of the queue.

**Figure 1.**
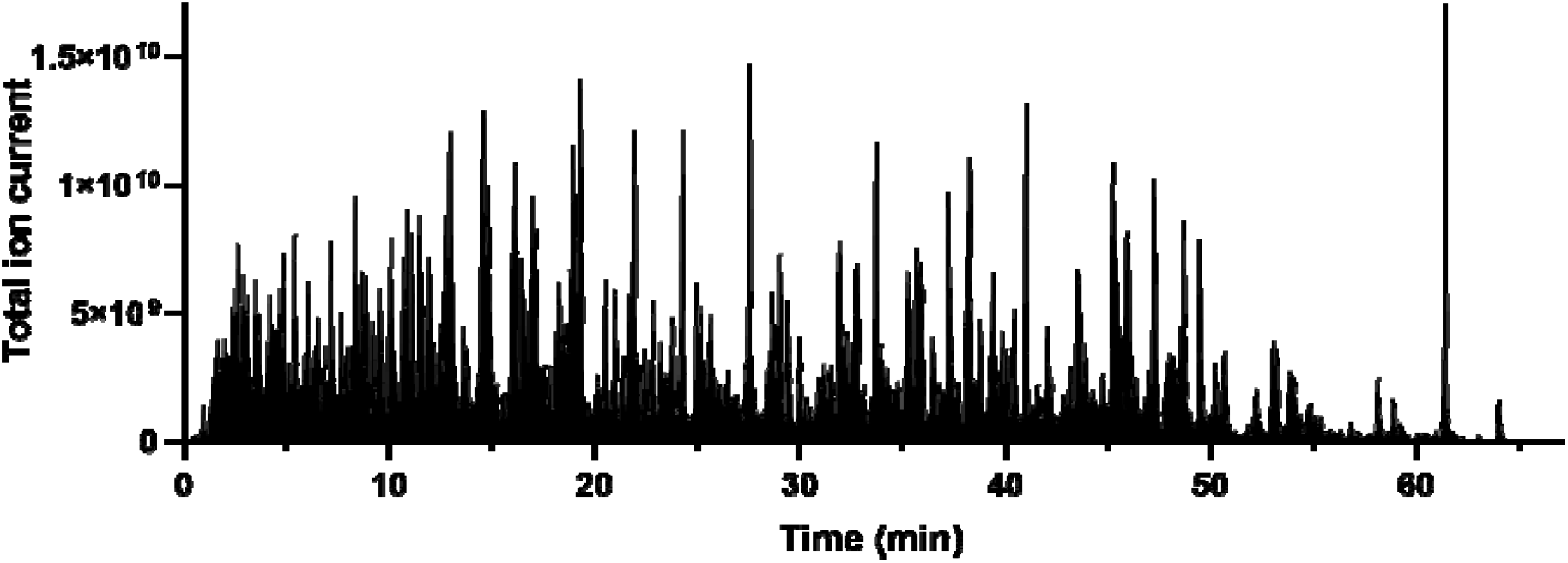
Representative chromatogram of tryptic digest of saliva. A tryptic digest (∼40 µg) was analyzed using microflow-LC-MS/MS and visualized as total ion current.

We achieved high proteome coverage of the saliva proteome (∼3,000) and high precision in a one-hour run. Database searching and quantification of de-multiplexed data was performed using Spectronaut. Based on previous studies of the saliva microbiome,^26^ we appended sequences for *Streptococcus mitis* (STRMT), *Gamella haemolysans* (9BACL) and *Granulicatella adiacens* (9LACT). Along with the 6 subject samples and 3 additional quality control samples, there were 3,550 total protein groups quantified from 36,547 unique peptide precursors, with 3,076 human protein groups, 332 *S. mitis* protein groups, 102 *G. haemolyans* protein groups and 42 *G. adiacens* protein groups (**Table S2 and S3**). Analytical precision was good, as measured by the percent coefficient of variation (%CV) of the three SPQC runs versus individuals, a mean %CV of 10% vs. 33%, respectively (**Table S3**). Hierarchical clustering suggested that individual biological variability was the major determinant of the saliva proteome, as sample clustered by subject rather than treatment (**Figure 2**).

**Figure 2.**
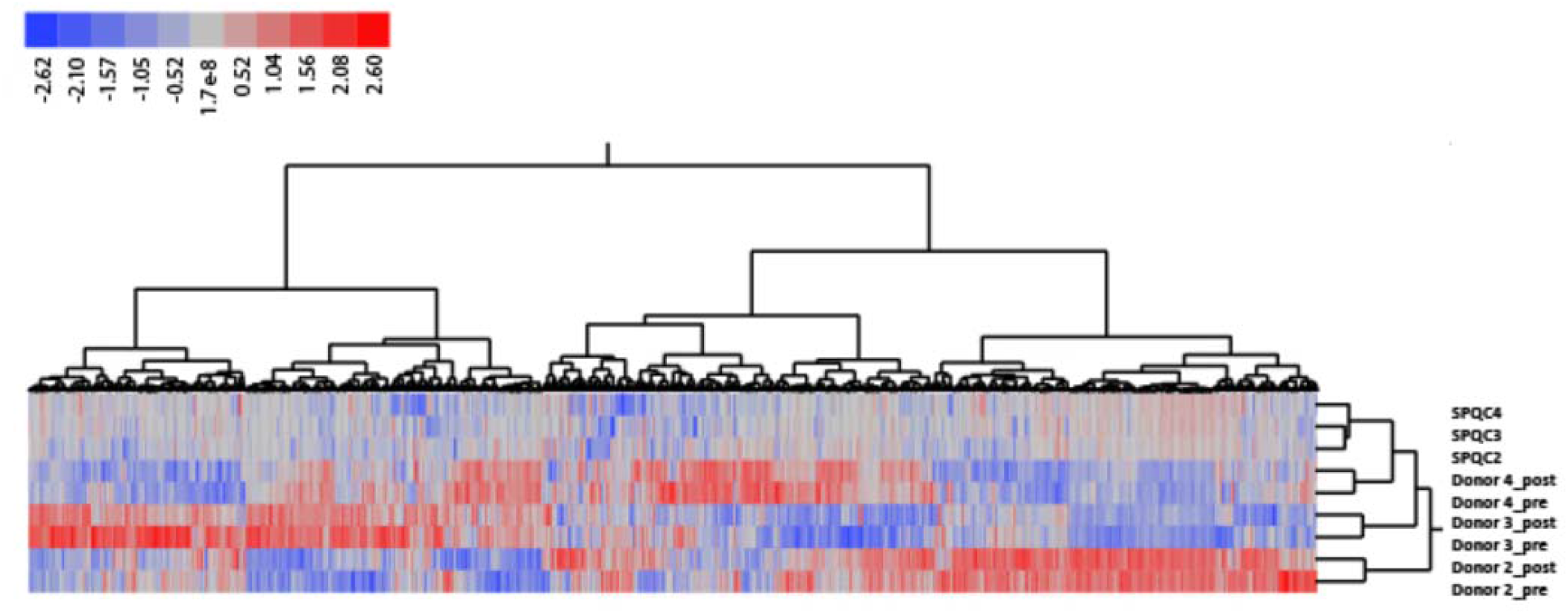
Heatmap of protein group expression. Z-scored abundances were exported from Spectronaut, and two-dimensional hierarchical clustering was performed in JMP Pro 16 using Ward’s method. Legend shows color scale of z-score values.

A paired statistical analysis was performed using the generalized linear model (glm) function in edgeR.^22,23^ There were and ∼300 proteins that had a p<0.05 between the pre-and post-octreotide groups ∼50 proteins with an FDR-corrected p<0.05 between pre-and post-groups (**Table S4**). These results were visualized using a volcano plot after filtering on proteins quantified by 2 more or unique precursors (**Figure 3).** Both human and bacterial proteins were among the proteins altered by octreotide treatment. Notably, four isoforms of the human cystatins, belonging to a family of cysteine proteases, had significantly lower abundance after treatment (**Figure 4**). Based on clustering (**Figure 2 and Table S4**), histatin-1 and (HIS1_HUMAN) and proline-rich protein 27 (PRR27_HUMAN), two other significantly downregulated proteins, had a similar expression pattern as three of the cystatins (CYTS; CYTD and CYTN).

**Figure 3.**
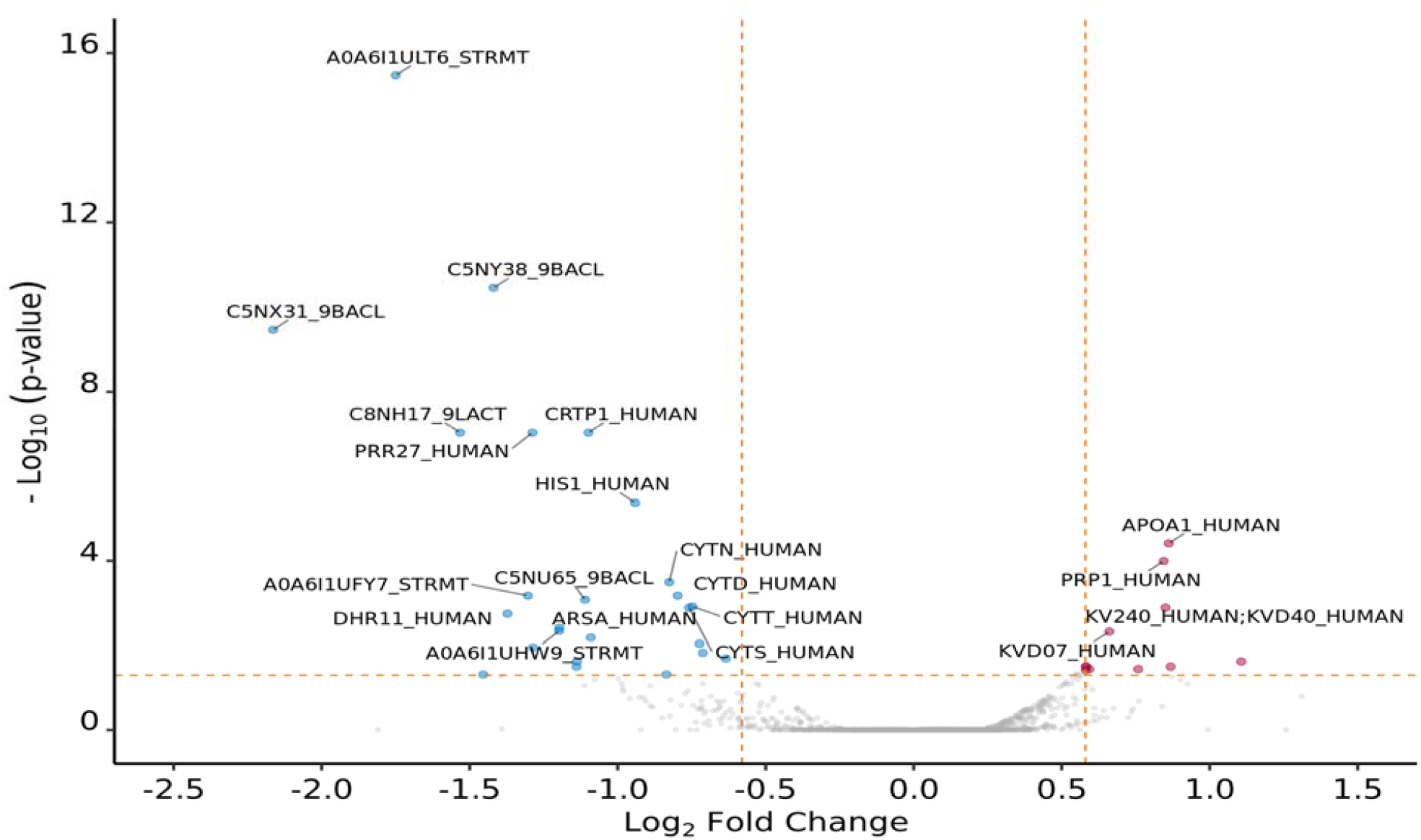
Volcano plot of proteins with differential abundance in saliva post-versus pre-octreotide w/ p-values. Statistical analysis used the generalized linear model (glm) function in edgeR. Log2FC and p-value data was filtered by proteins quantified by more than one precursor and visualized using ggVolcanoR.24 The top 20 proteins with absolute log2FC > 0.58 and FDR-corrected p-value <0.05 were labeled as significant.

**Figure 4.**
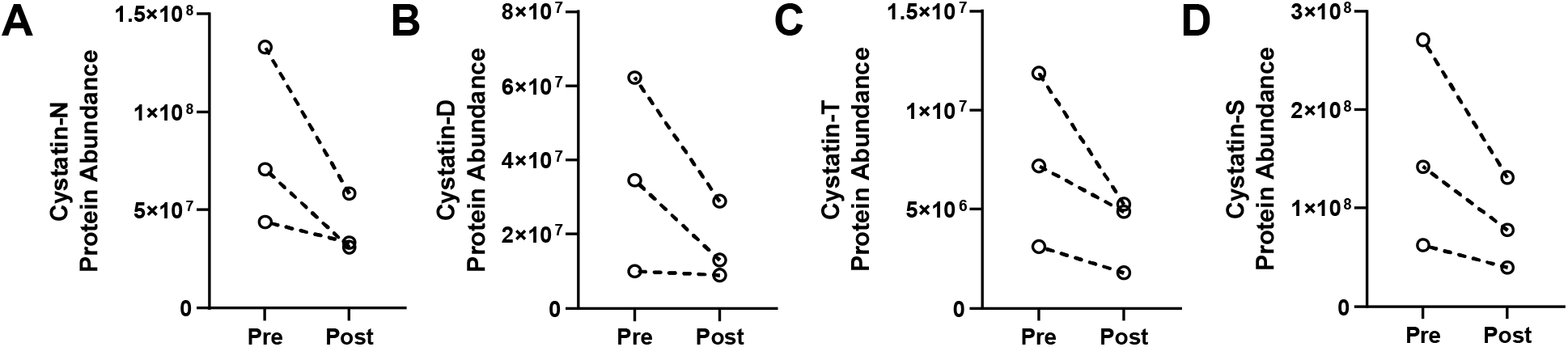
Human cystatin proteins with differential abundance. Protein abundances were plotted for the three subjects, with intraindividual pre-and post-octreotide values separated by a dotted line. Cystatin proteins which were significantly lower after octreotide treatment included (**A**) cystatin-SN (CYTN_HUMAN), (**B**) cystatin-D (CYTD_HUMAN), (**C**) cystatin-SA (CYTT_HUMAN) and (**D**) cystatin-(CYTS_HUMAN). Data were plotted in GraphPad Prism.

Rank protein abundance in saliva protein was estimated using the average MaxLFQ values of three SPQC samples (**Figure 5A**). Bacterial proteins were absent in the top two orders of magnitude, and the affected cystatin proteins were estimated to be among the 200 most abundant proteins (**Figure 5A**) but not within the top twenty (**Figure 5B**). Note that a similar analysis by Grassl et al.^27^ excluded keratins due the fact they are potential contaminants. However, levels of cytokeratins exhibited low intra-individual variability, and high inter-subject variability, suggesting that they are endogenous to saliva and not laboratory contaminants.

**Figure 5.**
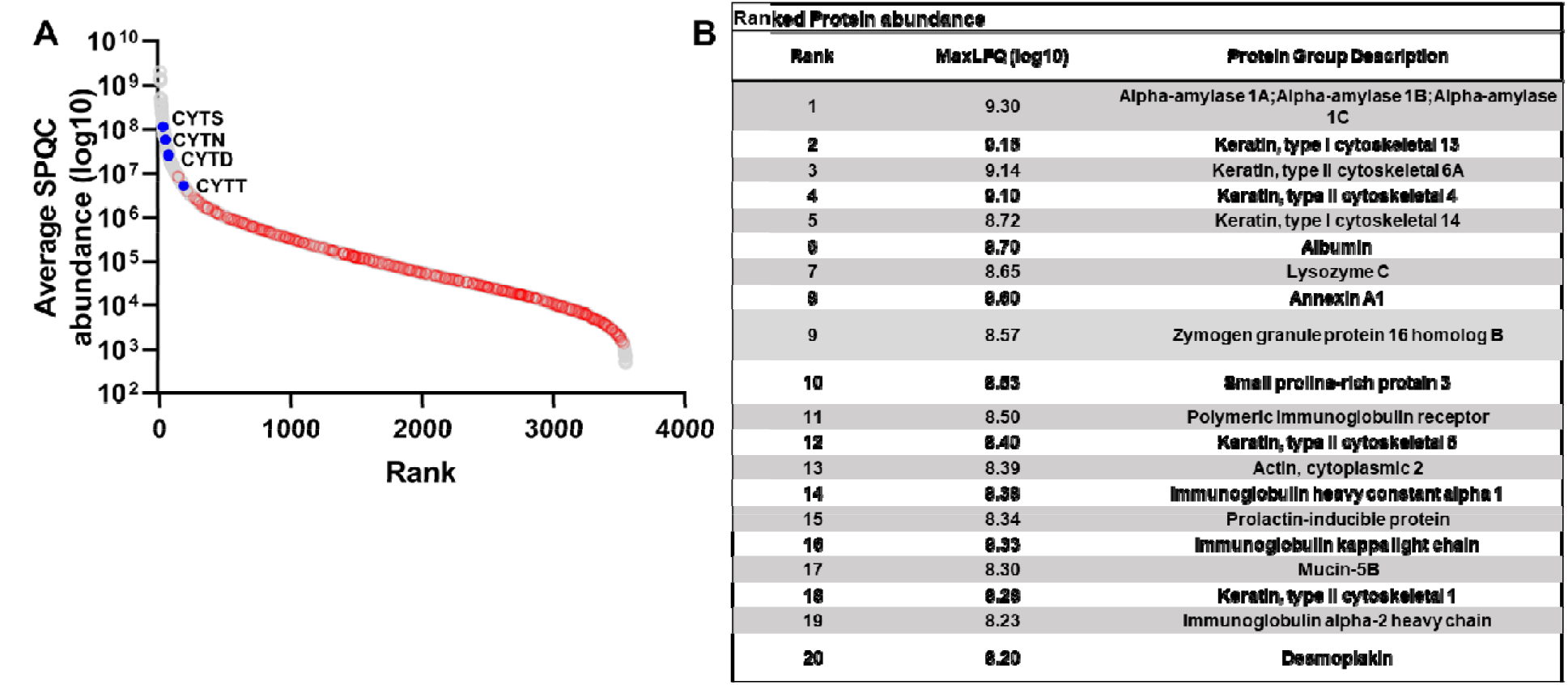
Ranked protein abundance in saliva. **(A)** Average protein abundances in SPQC runs (log10 MaxLFQ values) were plotted for all quantified proteins. grey, human proteins; red, *Streptococcus mitis* proteins; blue, cystatin proteins that were significantly lower after octreotide treatment. (B) Top twenty proteins based on rank abundance (excluding porcine trypsin; **Table S4**).

## DISCUSSION

This pilot study is the first to report octreotide induced salivary proteome alterations using mass spectrometry-based proteomics. Furthermore, our findings provide a potential mechanism the role of octreotide in wound healing. Microflow liquid chromatography coupled to data-independent acquisition quantified a comparable number of saliva proteins as compared to a recent nanoflowLC analysis^27^ in approximately one-half the total instrument cycle time. One class of proteins significantly downregulated were cystatins.

Cystatins belong to a superfamily of ancestrally related proteins, most of which are inhibitors of cysteine proteinases. The levels of salivary cystatins have been examined in the oral health literature, which has found a significant range in cystatin abundance amongst periodontally healthy individuals.^28,29^Cysteine proteinases include a number of cathepsins found in the oral environment. For example, cathepsin S is expressed in both microvascular endothelial cells as well as periodontal ligament cells. Shi and colleagues developed both in vivo and in vitro models demonstrated that deficiency in cathepsin S impairs microvascular angiogenesis in the context of wound healing.^30^ Furthermore, Memmert et. al utilized an in-vitro wound healing assay to show that cathepsin S significantly enhanced wound healing rates by increasing cell proliferation, migration, and wound closure. These authors further postulate that the role of cathepsins may be inhibited by periodontal flora in impairing wound healing.^31^

By downregulation of cystatins in the saliva, there is decreased inhibition of cysteine proteases such as Cathepsin S. This results in increased cysteine protease activity that has been linked to enhanced angiogenic response, cell proliferation and migration that have resulted in improved wound healing. These insights provide first steps at furthering our understanding of octreotide’s effects on saliva and reports of improved PCF healing.

This study is limited by its small sample size due to the COVID-19 restrictions that were imposed during the time of this study, as well as sample analysis-related costs. We were unfortunately forced to exclude one subject due to insufficient volume and protein concentration, so while the microflowLC approach is useful for throughput and robustness, it may suffer from sensitivity and is not preferred in sample-limited circumstances.

Another limitation of this study is that it does not evaluate the effects of drug in a disease population; rather, we can only infer potential mechanisms of action from the changes to control subject. Nonetheless, the relative abundance of approximately 50 proteins (of >3,500 quantified) were significantly altered after octreotide treatment, and cystatins appeared to be one of the major affected classes of host proteins.

In summary, this work represents the first to report octreotide induced alterations in saliva and a potential mechanism of action that could account for improved healing of PCF. The key finding is a significant decrease in cystatins that result in likely increased activity of cysteine proteinases, which has been associated with improved wound healing. Further research in a larger cohort is needed to investigate this potential mechanism of action as it relates to octreotide use and PCF closure.

## Data Availability

All data produced in the present study are available upon reasonable request to the authors

## Acknowledgments

We would like to acknowledge the clinical research support of Amy Walker, Research Program Lead, and Victoria Eifert, Senior Clinical Research Coordinator. We also thank Robert Plumb (Waters Corp) for supplying the custom-format ACQUITY Premier column used in this study.

## Supplemental Data

A supplemental file contains metadata (**Table S1**), normalized peptide (**Table S2**) and protein abundances, including analyses (**Table S3**).

## Notes

### Competing Interest Statement

The authors have declared no competing interest.

### Clinical Trial

NCT05340192

### Funding Statement

This study did not receive any funding

### Author Declarations

The IRB of Duke University gave ethical approval for this work.

